# SELF-POLICING COVID-19 AND CIVIC RESPONSIBILITIES IN LAGOS METROPOLIS, NIGERIA

**DOI:** 10.1101/2020.05.08.20092080

**Authors:** Ayomide Ilori

## Abstract

This study investigated self-policing COVID-19 and civic responsibilities in Lagos Metropolis, Nigeria adopting an online qualitative interview due to the current lockdown that denied field (face to face) interview. Fifty out of the feedbacks from the online interview were picked randomly to arrive at the conclusion of this study. The feedbacks suggested that there is adequate awareness of the COVID-19 pandemic among the people living in Lagos Metropolis, Nigeria and that they are following the directives of federal and state governments in an effort to reduce the community transmission of the infectious diseases. However, the ban on public gatherings and movements has made it impossible for many homes to meet their basic needs especially feeding. The government provided palliatives have also been largely insufficient to cater for the vulnerable. There could be a crisis (such as hunger) and the breakdown of law and order if the government does not increase their capacity to mitigate the hardship which the ongoing lockdown has imposed on the people.

## Introduction

COVID-19 has been traced to the unidentified pneumonia that started from Wuhan, China in December 2019. It is a severe respiratory syndrome in humans. According to Chang, Yan and Wang (2019), the World Health Organization (WHO) declared COVID-19 as an international public health emergency in 2019. However, aside the current pandemics of COVID-19, there were two earlier coronavirus infections-SARS in 2002-2003 and Middle East Respiratory Syndrome (MERS) in 2012 which caused severe respiratory syndrome in humans (Chang, Yan and Wang, 2020). In December 2019, some set of people at a seafood Market in Wuhan, China were diagnosed to have pneumonia. The cause of the illness could not be identified and was later labeled in 2019, the first set of the novel coronavirus (2019-1 nCov) case by WHO in January 7, 2020 (Huang, Wang, Li, Ren, Zhao and Hu, 2020). Subsequently the virus was renamed the Sever Acute Respiratory Syndrome Coronavirus 2 (SARS-CoV-2) and the infectious disease it causes was tagged Coronavirus Disease 2019 (COVID-19). Hence, on 30th January 2020, the WHO declared the disease as a global pandemic.

In efforts to mitigate the spread of the virus, many countries including Nigeria have used a combination of various measures such as restriction of movements, physical/social distancing, ban of public gatherings and place of worships, self-isolation and a 14-day quarantine period for returnees from some high risk countries. Others include aggressive campaign for good personal hygiene, regular hands washing with soap under running water, use of alcohol-based hand sanitizers and use of face masks. In addition, schools, companies and other public buildings were closed down to prevent community transmission of the disease. The President of Nigeria in a nationwide address on March 29, 2020 imposed a total lockdown in Ogun, Abuja and the country’s commercial capital Lagos, for an initial period of two weeks so that the Federal Ministry of Health and Nigerian Centre for Disease Control (NCDC) and other stakeholders in the public health sector can make available the resources needed to control the spread of the disease. According to the President, the 14-day initial period of lockdown will also be used to identify, trace and isolate all individuals that had come in contact with confirmed cases, ensure the treatment of confirmed cases and prevent further spread to other states.

However, prominent Nigerians like the Nobel Laureate, Prof Wole Soyinka and legal luminaries-Femi Falana and Ebun-Olu Adegboruwa, faulted 14-day lockdown directive of the President stating that the powers to impose a lockdown on any state of the federation should be with the backing of the National Assembly. This was however refuted by the presidency explaining that he acted in accordance with the provisions of the Quarantine Act of 1926. On Monday, March 30, 2020 the President signed the COVID-19 Federal Government regulation. The lockdown was later extended for 14 days on April 13, 2020 and another 7 days on April 27, 2020. However, in his most recent address on April 27, 2020, the President announced that there will be a gradual relaxation of the lockdown orders from May 2, 2020 while inter-state borders shall remain closed. As at 26 of April, 2020 according to the Nigeria Center for Diseases and Control (NCDC), Nigeria currently have 1273 total confirmed cases, 239 discharged and 40 deaths with 786 cases, 154 recovered and 22 deaths in Lagos Metropolis, Southwest region of the country. This study therefore investigates self-policing covid-19 and civic responsibilities in the Lagos Metropolis, Nigeria during these periods.

### COVID-19 in Nigeria and Government Designed Measures for Control

Nigeria recorded her index COVID-19 case on Feb 27, 2020. The index case was an Italian who had arrived in the country through the Lagos international airport two days earlier. He traveled from Lagos to his place of work-a factory in Ogun state where he became ill and was later confirmed to have been infected with the virus after he was returned to Lagos. Efforts were thereafter put in place by the two state governments to identify and trace the contacts of the index case. In less than two weeks, 27 suspected cases were identified across five states of Lagos, Edo, Abuja Ogun and Kano with two confirmed as positive cases. Similar to COVID-19, the first case of the Ebola epidemic in 2014 was also recorded in Lagos-the victim also came into the country through the same airport. The dense population of Lagos, its overstrained infrastructure, and the fact that it is a major regional transit hub for air, land, and sea transport created the perfect conditions for the spread of Ebola in Nigeria. Within weeks, nineteen (19) cases were recorded in two states of Lagos and Rivers State with eight (8) deaths (Musa, Nasidi, Shuaib and Nguku, 2016). Nevertheless, Nigeria’s aggressive and coordinated response successfully controlled the Ebola epidemic.

Due to its cosmopolitan nature and inadequate public health infrastructure, many fear that an outbreak of the disease in the country may be difficult to handle which is why stringent public health measures must be put in place. After it was declared as a global pandemic by the WHO, Nigeria was one of the first countries to identify the risk immediately plan response for COVID-19. To immediately curtail the increase of the virus, there was a national coordination and a multi-sectorial national coronavirus preparedness group was set up by the NCDC on January 7, 2020. Within a month, diagnostic capacity for COVID-19 as increased with the set up of three (3) laboratories in the country with the NCDC response team meeting daily to assess the risks to the nation and review its response to it.

In most of the states in the country, the primary measure put in place by the Governors was to ban interstate movements through the land, air or water followed by the closure of some government and private institutions, markets and other public places. However, when some of the states recorded their index cases, these measures were reviewed to include lockdown and curfew directives. Lagos and Ogun states seem to have taken more stringent measures to control the spread of the virus in their respective states by putting efforts together to increase their testing centers and isolation centers. For instance, the Lagos State governor under the State Public Health Law and the Federal Quarantine Act, Q2 LFN 2004, issued the Lagos State Infectious Diseases (Emergency Prevention) Regulations 2020 which designated COVID-19 as a dangerous infectious disease in the state. The regulations also classify its violation by anybody as a crime with violators liable to an option of fine or imprisonment as provided in the quarantine act and Lagos public health laws. On April 6, 2020, a popular actress-Funke Akindele and her husband were arraigned for hosting a house party which violated the provisions of the regulations. The court sentenced them to 14 days of community with a fine of two hundred thousand naira (N200, 000).

In a bid to keep the economy from total collapse and support business operations in the country, the President during his speech gave a moratorium of three months on loans implemented through the Bank of Industry, Bank of Agriculture and the Nigeria Export Import Bank. Financial and money markets were allowed to run skeletal operations so that the people can have access to online banking services. Senior or critical officers of some private and public institutions such as the Central Bank of Nigeria (CBN), deposit money banks, the Nigeria Interbank Settlement System (NIBSS), mobile money operators, payment solution providers were also exempted from the restriction of movement orders.

## Theoretical Framework

### The Health Belief Model (HBM)

This study adopts the Health Belief Model (HBM) to explain the behavioural change of Nigerians regarding the current Corona virus pandemic in the world, with thousands of infected persons in Nigeria and majority of the index cases in Lagos State. The HBM developed in the 1950’s, which has a psychological perspective likewise interactionism perspective on health and illness, is therefore considered adequate for this study. HBM holds that knowledge, prevention and control of COVID-19 are functions of individual beliefs. This model assumes that people must possess minimal level of health information and motivations in order to seek prevention or change health related behaviours during the current pandemic of COVID-19 and Post-COVID-19. For knowledge, prevention and control of COVID-19; hence; HBM is a decision making model that outlines, explains and predicts the likelihood of initiating certain health behaviours (Rosenstock, 1974). HBM is premised on key constructs bordering on factors that can promote knowledge, prevention and control of COVID-19 in Lagos Metropolis by initiating some certain health behaviours. The key constructs that was originally based on the first four constructs, applies to this study, thus:

**Perceived Susceptibility** - each individual has his/her own perception of likelihood of experiencing a condition that would adversely affect his or her health. In the case of knowledge, prevention and control of COVID-19 in Lagos Metropolis, aside the collective awareness of the infectious virus, individuals differ in their perception of susceptibility to a disease. However the level of susceptibility is at three levels, that is, high, moderate and low susceptibility. Individuals at the low end could deny the possibility of contracting COVID-19. Individuals at the moderate category could admit to a statistical possibility of susceptibility to the infection. While individuals at the extreme of susceptibility feel there is serious danger that they will be infected and thus obliged to preventive measures stipulated by the government and health authorities in Lagos State. **Perceived Severity** - this refers to the beliefs individual holds concerning the effects that COVID-19 infection would have on one’s state of affairs, if infected and thus obliged to preventive measures. These effects can be considered from the point of view of the seriousness of a health condition of the individual i.e immune system or old age and thus the kind of prevention practices the individual would engage in, as stipulated by the government and health authorities in Lagos State. The knowledge about the level of infection determines the way people perceive the severity of the virus and their susceptibility. In this light, such individual could engage in prevention and control practices. **Perceived Benefits –** this is the gains or wellbeing an individual expects for taking action toward the prevention and control of COVID-19 as outline by government and authorities. Such actions are taken after an individual has accepted the susceptibility and severity of the pandemic. **Perceived Barriers** - action towards prevention and control of COVID-19 could be difficult even though an individual may believe that the benefits to taking action are great, due to barriers like inconvenience use of face masks and expensiveness of hygiene like hand sanitizers, face masks, washing of hand with soap and running water in Nigeria. However, make a person take these action could be personal dispositions such as age, sex, marital status or personal enabling factors such as income, place of residence, transportation, occupation and education. **Cues to Action** - this are prompting engagement to prompt health behavior like Nigeria Center for Disease and Control COVID-19 prevention and control measures cues to action, expected of every an individual’s in Nigeria to adhered to or forced to act. Such cues to action could be internal or external i.e the COVID-19 physiological symptoms and information of prevention and control from the health care provider, the media and significant others. However it requires a cue to action for the desired behaviour to occur. **Self-efficacy** - This refers to the level of a person’s confidence in his or her ability to successfully perform preventive and control measures of COVID-19 in Lagos metropolis as stipulated by the government and health authorities in Lagos State.

In sum, the HBM explains that the desire to prevent and control COVID-19 in Lagos Metropolis, Nigeria, relies on the level of knowing of individual responsibility to curtail the widespread of COVID-19 which are the lockdown in Lagos State, the popular physical/social distancing and personal hygiene like washing of hand with soap, coughing or sneezing on the elbow, use of hand sanitizers, face mask in public places and hand gloves and other measures that would prevent and control the widespread of COVID-19 in Lagos Metropolis, Nigeria.

## Research Setting and Study Population

The study was conducted in Lagos Metropolis, South-West Nigeria. The rationale behind the selection of Lagos for this study is due to the fact that the city has the highest index cases of COVID-19 in Nigeria. Modern-day Lagos, known as “Metropolitan Lagos”, and officially as “Lagos Metropolitan Area (1996) is an urban agglomeration or conurbation (Carpio, 2012) consisting of 16 LGAs including Ikeja, the state capital of Lagos State. This conurbation makes up 37% of Lagos State’s total land area, but accommodates about 85% of the state’s total population (Eko, 2012). The exact population of Metropolitan Lagos is disputed. In the 2006 federal census data, the conurbation had a population of about 8 million people (Lagos State Government, 2015). However, the figure was disputed by the Lagos State Government, which later released its own population data, putting the population of Lagos Metropolitan Area at approximately 16 million (Lagos State Government, 2015).As of 2015, unofficial figures put the population of “Greater Metropolitan Lagos”, which includes Lagos and its surrounding metro area, extending as far as into Ogun State, at approximately 21 million (Lagos State Government, 2015). However, residents of Lagos metropolis, constituted the population for this study.

## Methods

This study was exploratory and cross-sectional in design. Since it was impossible to administer interviews physically to participants, an online interview approach which is also qualitative in nature was employed to achieve the urgency that this study demands, during the COVID-19 pandemic that prompts lockdown and physical/social distancing. Ethical approval was obtained from the Faculty of Social Sciences Ethical Board, University of Ibadan. Hence, participation in the study was consensual, voluntary, and anonymous and the study poses no risk to the participants. However, this study used 50 randomly selected responses from the 96 respondents that filled the online interview, during the 14-days lockdown directive of the President of Nigeria. The data generated from the online interview went through careful transcription, detailed description, and interpretation of the emerging themes from the online interview. All categories of responses extracted online were compared and merged to draw out and/or create a clear picture of the emerging COVID-19 themes to further enhance the robustness of the findings and interpretations.

## Results and Discussion

This section discusses the results that emanated from the three objectives covered in this study. The findings were thematically presented to ensure logicality and lucidity of discourse with literature and theory reviewed for this study.

## Knowledge of Covid-19

As earlier established, COVID-19 was declared a pandemic and public health emergency of international concern (PHEIC) by the World Health Organization (WHO) on January 30, 2020 (Euro Surveillance, 2020). Currently, there is an increasing number of reported cases with a total of 2, 992, 501 confirmed cases, 877,254 recoveries and 206,878 deaths as at 26 of April, 2020 globally. In Nigeria as at April 26, 2020 according to available data from the Nigeria Center for Diseases and Control (NCDC) there are 1273 confirmed cases with 239 discharged and 40 deaths recorded. The Lagos Metropolis, Southwest region of Nigeria appears to be badly hit by the spread of the virus with 786 confirmed cases, 154 recoveries and 22 deaths. COVID-19 virus is primarily transmitted among people through respiratory droplets. According to WHO (2020), the COVID-19 is spread by human-to-human through droplets, feco-oral, and direct contact, with an incubation period of 2-14 days. This therefore informs this study to test the knowledge of COVID-19 transmission and establish the vulnerability level of the people to an infection. One of the participants asserted thus;

> Physical touch of infected person and contact with an infected place and later touching either of mouth, eye or ear. Also inhaling droplets. (Male/Married/Graduate Degree/Students/Lagos Metropolis, Nigeria)

In the word of another participant;

> Contact with infected patient through exposure to mucus or saliva of infected person(s) when the virus comes in contact with my eyes, nose or mouth. (Male/Single/Graduate Degree/Unemployed/Lagos Metropolis, Nigeria)

This participant also asserted thus:

> Through touching of surfaces infected by a carrier and having contact with an infected person and touching of eyes, nose or mouth with an infected hand. (Male/Married/Masters/Employed/Lagos Metropolis, Nigeria)

In a similar response, a participant stated that:

> By coming in contact with the virus when it’s on a surface through our hands and also when the fluid of an infected person gets to the face or into the mouth. (Female/Married/ Graduate Degree/Students/Lagos Metropolis, Nigeria)

In the words of another participant:

> Physical contact with an infected person or surface, then transfer it to the body through the eyes, nose or mouth. (Female/Married/ Graduate Degree/Unemployed/Lagos Metropolis, Nigeria)

This participant also said that:

> By touching surfaces, body contact and fluid from shaking of hands and cough that are already contaminated with the virus and later touching your mouth, nose eyes etc. (Female/Married/ Graduate Degree/Employed/Lagos Metropolis, Nigeria)

It can be deduced that the residents of Lagos Metropolis are aware that COVID-19 is infectious. This can be attributed to the awareness efforts of the government, health authorities, individuals and corporate bodies in educating people about the virus. Such awareness according to the Health Belief Model, informs individual susceptibility to the virus and such informed knowledge becomes internalized and it becomes a collective consciousness to prevent and control the widespread of the infectious diseases in Lagos Metropolis. Furthermore, the symptom of COVID-19 infection was identified as responses showed that residents in Lagos Metropolis are fully aware of the symptoms of COVID-19 infection which indicates that the governments through the Nigeria Center for Diseases and Control (NCDC) and other health agencies have adequately invested in sensitization campaigns to inform Nigerians about the symptoms of the virus. According to one of the participants:

> Dry cough, running nose, sneezing, and breathing problem. (Male/Single/HND/Employed/Lagos Metropolis, Nigeria)

In the word of another participant:

> Fever, difficulty in breathing, sore throat, dry coughing. (Female/Single/Graduate Degree/Unemployed/Lagos Metropolis, Nigeria)

Another participant identified the symptom as;

> Fever, difficulty in breathing and Body weakness. (Female/Single/Graduate Degree/Employed/Lagos Metropolis, Nigeria)

In a similar response:

> Persistent cough, Headache, shortness of breath, fever. (Male/Single/Graduate Degree/Employed/Lagos Metropolis, Nigeria).

Another participant stated that:

> Throat itch, cough, Sneezing, breathing difficulty, fever, among others. (Female/Single/Graduate Degree/Employed/Lagos Metropolis, Nigeria)

The above narrative indicates that the people in Lagos Metropolis are aware of the symptoms of COVID-19 infection. This shows how effective the awareness of COVID 19 in Nigeria where Lagos Metropolis is the commercial center of the country has become. Notably also, the WHO embarked on several online training sessions and materials on COVID-19 in various languages to strengthen the preventive strategies, including raising awareness, and training healthcare worker’s preparedness activities (WHO, 2020). In purview of the embarked upon awareness about COVID-19 in the country, it was also affirmed from the narratives of the participants’ that there is possibility reinfection of COVID-19. One of the participants asserted thus:

> Yes, coming in contact with an infected person. (Male/Single/ Graduate degree/Employed/Lagos Metropolis, Nigeria)

Another participant stated otherwise:

> Yes, if a treated person does not maintain good personal hygiene. (Male/Married/ Graduate degree /Employed/Lagos Metropolis, Nigeria)

In another affirmative response, it was stated thus:

> Yes, if the safety measures are not observed. I read some people get re-infected. (Male/Single/ Degree/Employed/Lagos Metropolis, Nigeria)

In a similar response, another participant asserted thus:

> Only if the person is careful, there is possibility of reinfection. (Female/Single/ Graduate Degree/Unemployed/Lagos Metropolis, Nigeria)

Another participant put it that:

> It has been stated that there is a possibility of reinfection (Female/Single/ Graduate Degree/Student/Lagos Metropolis, Nigeria)

From the above submission therefore, COVID-19 is an infectious disease that has become known regarding the scientific knowledge of the infectious diseases-infection, symptoms and reinfection. Hence, global health experts and African governments have shown concern about the spread of COVID-19 and potential for more than 2 million deaths in sub-Saharan Africa if no action is taken (Walker, Whittaker and Watser, 2020).

### Prevention and Control of Covid-19

Due to the directive given by the President and other states governor to curtail the epidemic of COVID-19 in Nigeria especially interstate, intrastate and community transmission, Nigerians readiness to take up responsibilities as directed by the president and respective state governors in South West to control the widespread of the infectious disease trough social distancing, personal hygiene like hand washing with soap and use of sanitizer was tested. According to Dahab, Zandvoort, Flasche, Warsame, Spiegel, Waldman and Checchi (2020), implementing personal hygiene and public health behaviors are necessary to curb the spread of coronavirus, such as hand washing and social distancing. Without sustained bans on large gatherings (including specific cultural and faith practices such as mass prayer gatherings, large weddings and funerals), these may create super-spreading events that accelerate transmission (Wong, Liu, Liu, Zhou, Bi, and Gao, 2015). However, the participants expressed readiness to curtail the further spread of the virus in Lagos metropolis.

In the words of one of the participant, the government directive was supported but there is need for palliatives as noted:

> Good idea but the government should have help cushion hunger by distributing relieve materials such as food and hand sanitizers. The curfew is appropriate to limit the spread of the pandemic from person to person. However, it will make life difficult for those who live on daily earnings. (Female/Married/ Graduate Degree/Employed/Lagos Metropolis, Nigeria)

Another online participant’s shows approval for the directive but also laid some important complain as thus:

> Well, it is not a bad idea but it is a bad idea for this government. Reasons being that, they never have anything under control, how can they put a curfew when people will starve? Provisions should be made and they should adequately monitor it so to ensure it gets to people and not ask them to gather themselves for it to aid spreading the disease like the case of Lagos State. No checkmated measure to ensure stability in this trying times. (Female/Single/ Graduate Degree/Unemployed/Lagos Metropolis, Nigeria)

In another similar response:

> It’s quite unreasonable unless the government provides other means of survival for the people. Measures should have been taken ahead of the curfew to cater for the population who feeds on daily earnings. (Female/Single/ Graduate Degree/Employed/Lagos Metropolis, Nigeria)

In the word of another participant:

> The lockdown and curfew to prevent and control COVID-19 is good but let d FG provides palliatives for the public to obey the stay at home order. (Female/Single/ Graduate Degree/Student/Lagos Metropolis, Nigeria)

In another similar response:

> The lockdown down by the government is needed, to help curb the pandemic until a cure is found. Best measure to avoid community spread is the lockdown directive by the government which is the same measure in other countries. (Male/Single/Married/ Graduate Degree/Student/Lagos Metropolis, Nigeria)

It can be deduced from the above responses, that residents of Lagos Metropolis, Nigeria are ready to prevent and control the widespread of COVID-19 in accordance with the government directive but there is need for palliatives to be provided by the government, to cushion the effect of hunger, as the current lockdown do not support daily means of survival. However, to guarantee the final success of the prevention and control of the infectious disease, HBM holds that perceived barriers to action like inconvenience use of face masks and expensiveness of hygiene like hand sanitizers, face masks, washing of hand with soap and running water in Nigeria and most especially huger palliatives, should be provided for people’s to comply to the established rules of behaviour to curtail widespread of COVID-19 in Lagos Metropolis, Nigeria. Furthermore, compliance to civic orders after COVID-19 was tested to ascertain the readiness to comply with expected civic regulation to curtail post COVID-19 infections. One of the participants submitted that:

> It depends on the individual if they are willing to comply. Some may do whatever they want regardless. (Female/Single/ Graduate Degree/Employed/Lagos Metropolis, Nigeria)

In a similar response, the readiness for future compliance to civic orders after COVID-19 was asserted thus:

> It depends on level of orientation. (Male/Single/ Graduate Degree/Employed/Lagos Metropolis, Nigeria)

Another participant corroborated thus:

> Certainly not everyone will comply with such civic order to curtail post COVID-19 infections. It depends on individual (Male/Single/ Graduate Degree/Student/Lagos Metropolis, Nigeria)

In the words of another participant, he expressed that future compliance to post COVID-19 civic orders is not certain:

> In the case in Nigeria, our people are stubborn unless you force them. (Male/Single/ Graduate Degree/Student /Lagos Metropolis, Nigeria)

Another participant asserted that:

> It depends on the situation at hand, future times (Female/Single/ Graduate Degree/Unemployed /Lagos Metropolis, Nigeria)

In the word of another participant, readiness for future compliance to civic order was ascertained:

> Yes. I do that always and will continue to do so. I have always obey government orders. (Female/Married/ Graduate Degree/Employed /Lagos Metropolis, Nigeria)

It can be deduced that the above responses suggest that noncompliance to post COVID-19 depends on the level of susceptibility to the infectious virus, its severity and cues to action, according to HBM. Also, compliance to post COVID-19 prevention and control in Lagos Metropolis depends on perceived benefits and absences of perceived barrier to action for COVID-19 prevention and control in future times.

### Covid-19 Civic Obligations

According to the quarantine and NCDC acts of the Nigeria federal government, it is expected that everyone complies with the government directives that are lockdown, social distancing and hygiene to curtail the spread of COVID-19. The setting up of mobile court to try offenders also indicates the readiness of the government to ensure civic obedience to such directives. This has recorded some success in the region. However, the participants agreed that COVID-19 14-day ‘lock down” directives is crucial to stop the spread of virus. One of the participants’ submitted that:

> Yes, the lock down poses a threat to my movement but have no choice than to stay safe. (Male/Single/ Graduate Degree/Student/Lagos Metropolis, Nigeria)

Another participant asserted otherwise:

> It’s just a civic responsibility during this pandemic. I do not see it as threat to my freedom, but a strategy to protect my family and I. (Female/Married/ Graduate Degree/Employed/Lagos Metropolis, Nigeria)

In the words of another participant:

> No, the measures are necessary for me. It’s just bored staying indoor. (Male/Married/ Graduate Degree/Student/Lagos Metropolis, Nigeria)

Another participant succinctly put it that:

> Yes, ooo, as an artiste…staying indoor not helpful this time. (Male/Married/ Graduate Degree/Student/Lagos Metropolis, Nigeria)

Another participant claimed that:

> No its not, we are safer at home than out in the streets and it is necessary for the purpose of preventing the spread of the virus. (Female/Married/ Graduate Degree/Employed/Lagos Metropolis, Nigeria)

In the words of another participants

> I do not see it as a threat. It is just a civic responsibility and a measure that is necessary for safety during this COVID-19 pandemic. (Female/Married/ Graduate Degree/Employed/Lagos Metropolis, Nigeria)

It can be deduced from the responses that the federal government and state government lock down is seen as a civic obligation that is necessary to curtail the widespread of COVID-19, regarding the Regulations signed 30th of March, 2020. In the regulations, the President directed the cessation of movements in Lagos, Ogun and FCT for an initial period of 14 days with effect from 11pm on Monday 30 March 2020. Citizens are to stay in their homes and all businesses and offices should be fully closed while travel to other states should be postponed. Furthermore, the participants identified what the restricted movement has done to their daily lives and living. A participant submitted that:

> Going around to work and general movement. Not being able to shop freely, going into town, church, and visiting my friends and family. (Female/Single/Graduate Degree/Unemployed/Lagos Metropolis, Nigeria)

Similarly, another participant responded as follow:

> Movement, going to work, moving freely from one location to another. But if staying indoors is the only way to avoid spread, then I’m happy to comply. (Female/single/graduate degree/unemployed/Lagos Metropolis, Nigeria)

Another participant who is a student put it thus:

> It has affected my going to school and some side hustling as a student. (Male/Single/Graduate Degree/Student/Lagos Metropolis, Nigeria)

In the words of another participant:

> Going about for my normal day to day hustle, freedom of movement and freedom to mingle with friends and so on. For now, this is needed. Desperate times, desperate measures. (Female/Single/Graduate Degree/Student/Lagos Metropolis, Nigeria)

In a very definite response, it was asserted thus:

> Use of health facility for antenatal care. (Female/Married/Graduate Degree/Student/Lagos Metropolis, Nigeria)

In the words of another online participant:

> My Search for job and Freedom of movement section 41 CFRN 1999. (Male/Single/Graduate Degree/Unemployed/Lagos Metropolis, Nigeria)

These above responses therefore affirm the compliance to the federal government and state government directives to curtail the widespread of COVID-19 in Lagos metropolis, Nigeria. However, in accordance with this government directives, HBM holds that, there should be cues to action from government and health authorities that will build the resident self-efficacy to successfully prevent and control spread of COVID-19 in Lagos metropolis, Nigeria. The government and health authorities directive has thus established a standard of behavior for residents in Lagos metropolis, Nigeria which include a variety of sanctions such arrest of offenders, public shame to enforce compliance to curtail the widespread of the infectious diseases due to the higher level of susceptibility to the virus and fear of community transmission of COVID-19 in Lagos metropolis.

## Conclusion

This study has shown that COVID-19 is generally known among Lagos Metropolis, Nigerian as an infectious disease and due to the threat it poses to social organization and human relationship therein, the government has put in place measures to curtail the widespread of the infectious virus which are lockdown, physical/social distancing, washing of hand with soap and water, hand sanitizers, use of face mask and hand gloves. The envisage spread and health damages that could submerge the capacity of the public health care system have encouraged the people to comply with federal and state government regulations and directives. This study therefore suggests that as residents of Lagos Metropolis, Nigeria have complied with the directive in accordance to their civic responsibilities, the government should avoid a “rebellion of sort” by providing necessary support to the people. The government should increase capacity to meet the yearnings of the people in providing more palliatives to tackle hunger and invest more in traditional list of immediate “basic needs” which in Nigeria are food (including water), shelter and electricity. This will further help maintain and guarantee the further compliance to COVID-19 prevention and control measures in Lagos metropolis, Nigeria that currently has the highest index case in the country. In addition, investing in basic needs, it will promote the obedience of civic order that is to curtail the widespread of COVID-19 in Lagos metropolis, Nigeria without accountability or reference to an external authority or use of force to compel obedience for residents.

## Data Availability

The data is available as extracted from the online interview

